# Digital Health Intervention to Promote Lifelong Specialized Care in Adults with Congenital Heart Disease: Study Design

**DOI:** 10.1101/2025.03.31.25324723

**Authors:** Anushree Agarwal, Joseph Valente, Karina Buenrostro, Katelyn Macholl, Juhi Mehta, Keerthana Reddy, Karina Manayan, Parang Kim, Aleah Sparks, Kunyi Li, Pranav Ahuja, Kevin Sun, Kimberly Payton, Mark D. Norris, Katia Bravo-Jaimes, Leigh Reardon, Philip Moons, Megumi J. Okumura, Gregory M. Marcus, Michelle Gurvitz

**Author notes:** Address for correspondence: Anushree Agarwal, MD, MAS Associate Professor of Medicine, Adult Congenital Heart Disease Section, Division of Cardiology University of California San Francisco 500 Parnassus Avenue, M-1177B, Box 0124 San Francisco, CA 94143-0124.

## Abstract

**Background:** Adults with congenital heart disease (ACHD) frequently experience gaps in lifelong ACHD specialty care, leading to preventable complications, hospitalizations, and premature mortality. However, effective, scalable, accessible, and sustainable strategies to reduce these gaps are lacking. Digital health offers potential solutions but requires a rigorous scientific approach in its design to address the needs of the target population.

**Objective:** To describe the development of a theory-based, community co-designed digital health intervention to improve lifelong ACHD care.

**Methods:** We integrated theory-based behavioral frameworks, semi-structured qualitative interviews with patients and clinicians, and a community-engaged approach to develop a digital health intervention for ACHD. The primary behavioral target was completing an ACHD specialist appointment. We conducted Capability, Opportunity, Motivation, and Behavior (COM-B)-guided semi-structured interviews with ACHD patients and clinicians to identify barriers to specialized care amenable to a digital intervention and patient-centered goals for the digital tool. The community partners helped develop key intervention objectives, create a theory-driven framework, and specify how each intervention component targets specific COM-B barriers.

**Results:** We interviewed 54 participants (37 ACHD patients and 17 clinicians) and engaged 21 community partners representing 4 advocacy organizations. Design objectives emphasized addressing patient loneliness, ensuring accessibility and credibility, and enabling scalability while centering patient perspectives. Participants identified four priorities: providing credible resources, uplifting patient voices, customizing to patient needs, and centering positivity and joy. The digital tool, named by community partners as Empower My Congenital Heart (EMCH), was designed within the web- and mobile-based, Apple- and Android-compatible, Eureka Digital Research platform (University of California, San Francisco). Key intervention components included educational modules, peer support, appointment planning nudges, and a digital medical passport. The EMCH’s theory-driven framework specifies how each intervention component targets specific COM-B barriers to specialized ACHD care.

**Conclusions:** The theory-driven, community co-designed EMCH digital tool is a rapidly scalable approach to improve guideline-recommended specialized ACHD care. Future studies will evaluate the acceptability, feasibility, effectiveness, and implementation outcomes to provide insight into a novel tool to reduce preventable morbidity and mortality in ACHD.

**Trial Registration:** The National Clinical Trial number for this study is NCT06581484.

## Introduction

Adults with congenital heart disease (ACHD) are a rapidly growing population [1,2]. The American Heart Association and American College of Cardiology guidelines provide recommendations on the frequency and intervals at which ACHD patients should receive lifelong care from clinicians specialized in managing ACHD [3,4]. Despite this, up to 85% of ACHD patients experience gaps in receiving specialized care throughout their adult life [5–8]. Those with care gaps are at a higher risk of poor outcomes, including emergent admissions, need for urgent cardiac procedures, and mortality [9–11]. Thus, there is a need to identify accessible, scalable, and sustainable strategies to reduce the care gaps and improve outcomes for ACHD patients.

Individual-, provider-, and system-level barriers contribute to gaps in lifelong ACHD care [5,12,13]. Interventions, such as education and support for navigating the health system, can reduce gaps [14–16]. However, existing interventions are clinic-based (nurse-led education, transition clinics); limited to single-center studies of 16-21-year-olds focused on transfer from pediatric to ACHD care; and are resource-intensive, inconsistently implemented, and constrained by reimbursement, clinic time, and missed visits [17–19]. Effective and scalable strategies to support all adults (≥18 years), including those who did not transfer from pediatrics or were later lost from care, to both establish and maintain life-long specialized ACHD care remain unknown. Web- and mobile-based solutions offer a scalable way to overcome these barriers by engaging patients outside of clinic visits [20,21]. Although 94% of ACHD patients use smartphones and most report openness to app-based solutions [22], these tools have not yet been developed to promote lifelong specialized ACHD care.

App-based interventions to improve guideline-adherent care need to be grounded in evidence-based theoretical frameworks [21,23]. This results in interventions being more robust, adaptable, and effective in achieving lasting behavior change in real-world settings, not just in ideal research environments [24,25]. Also, designing interventions to change behaviors requires an understanding of the perspective and psychosocial context of the people who will use them - a concept fundamental to community-engaged research [26–28]. Combining theory- and community-driven approaches is vital to ensure that interventions are usable, acceptable, and move beyond *what* works to *how* to make it work in diverse, complex environments [25,29]. To our knowledge, no existing ACHD interventions, digital or otherwise, have employed this combined theory- and community-driven approach.

Thus, the aim of this paper is to describe a theory- and community-driven approach to designing a digital health intervention to promote lifelong ACHD specialist care. The methods and findings can inform others developing interventions to improve care for ACHD and other chronic lifelong conditions.

## Methods

### Overview of the intervention development process

We began by establishing guiding principles for intervention design (**Table 1**). We then integrated behavioral frameworks with semi-structured interviews and community-engaged research to identify and refine intervention components for promoting lifelong specialized ACHD care (**Figure 1**).

**Figure 1:**
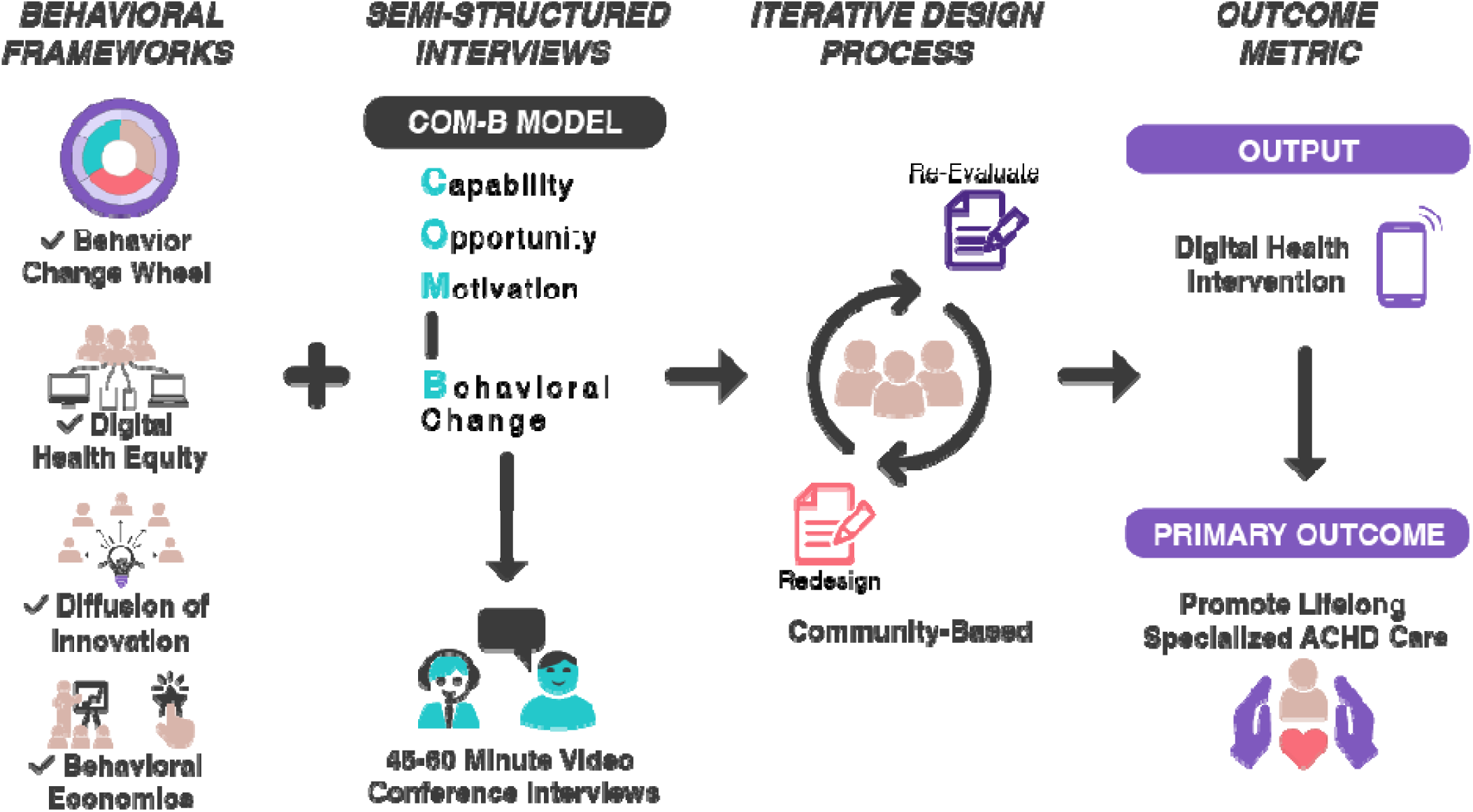
Overview of the Digital Health Intervention Planning and Development Process. Behavioral frameworks (Behavior Change Wheel, Digital Health Equity, Diffusion of Innovation, and Behavioral Economics) were integrated with semi-structured interviews and an iterative design process with the community partners. This process optimized the development of a digital health intervention with key objectives to build patient activation and empowerment skills and promote lifelong adult congenital heart disease specialized (ACHD) care.

**Table 1.**
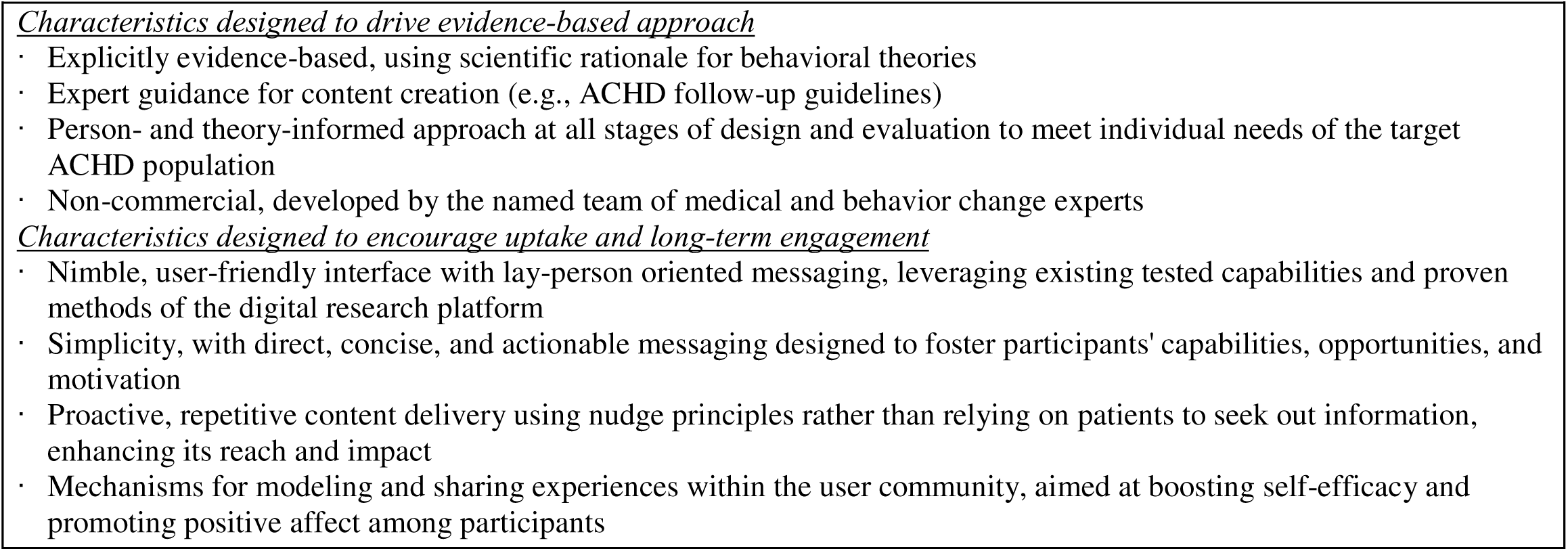
Guiding Philosophy for the Design of an ACHD Digital Health Intervention.

#### Behavioral frameworks

We selected evidence-based behavioral frameworks to guide intervention development. The study team chose 4 primary frameworks as they complemented the needed areas to design the digital tool. First, the Behavior Change Wheel (BCW) theoretical model was primarily used to provide a rational, evidence-informed approach to specify the target behavior, identify the barriers and enablers of the behavior, and select the means by which intervention can change behaviors [30,31]. To make the intervention equitable and relevant to diverse ACHD patients, we incorporated the principles of digital health equity, diffusion of innovation, and behavioral economics in its design and adoption (**Table 2**) [32–34].

**Table 2.** Theoretical frameworks for designing ACHD digital intervention.

|  |  |
| --- | --- |
| Frameworks | Summary of the Theoretical Frameworks |
| Behavioral Change Wheel (BCW) [30] | 1. Define health problem in behavioral terms (e.g., scheduling and completing timely ACHD specialist visit)<br>2. Understand determinants of behavior (Capability, Opportunity, Motivation for Behavior Change-COM-B Model)<br>3. Link determinants of behavior to identify domains most likely to influence behavior<br>4. Select intervention functions that are means by which an intervention changes behavior (i.e. education, enablement, persuasion, or training) |
| Digital Health Equity [32] | Apply concepts of:<br>1. Health literacy: include relevant content: assume no background knowledge of participants, avoid cognitive burden from too much information and use simple numbers and percentages to develop content<br>2. Readability: use plain and clear language tested with target population<br>3. Ease of Use: accessible on various platforms, format conducive to comprehension; content appeals to users of different identities and backgrounds |
| Diffusion of Innovation [33] | Engaging all levels of adopters (highly engaged and less engaged CHD patients) and CHD champions in the design, adoption, and dissemination of the intervention |
| Behavioral Economics [34] | Nudges and default settings of the digital tool alter behavior and facilitate decision-making process. |

The BCW framework is based on multiple models of health behavior and is designed to enable the systematic development of interventions. The BCW has been identified as an effective framework at changing individual health behavior in various cardiac and noncardiac chronic conditions but has not yet been used to promote lifelong ACHD care [35–40]. The core driver of behavior in BCW (or hub of the wheel) is the COM-B model, which consists of three necessary conditions for a given ‘**B**ehavior’ to occur: (1) ‘**C**apability’ (psychological/physical); (2) ‘**O**pportunity’ (physical/social); and (3) ‘**M**otivation’ (reflective/automatic). Exploration of a given behavior in relation to the COM-B components helped identify which psychological determinants need to be addressed to achieve behavior change. The BCW framework then supports the selection of intervention functions and policy categories. Intervention function refers to broad categories of ways an intervention can change a behavior. As applicable, one or more of the intervention functions can be chosen and applied depending on the COM-B component targeted for change. The nine intervention functions described in BCW include education, persuasion, incentivization, coercion, training, restrictions, environmental restructuring, modeling, and enablement. Policy categories comprise the final outer layer, or the wheel’s rim, and help identify the types of policy categories one may wish to consider to further influence the drivers of behaviors (COM-B).

#### Semi-structured Interviews

The semi-structured interview for this study were part of a larger qualitative study that was conducted to identify barriers and enablers for lifelong specialized ACHD care, described in detail separately [12]. For this study, we used COM-B as a guide to identify barriers amenable to a digital intervention and the patient-centered goals for the digital tool. Briefly, we recruited ACHD patients (≥18 years) who spoke English or Spanish and could provide informed consent. Initially, patients were recruited from University of California San Francisco (UCSF) using chart review and purposefully sampled [41] to recruit those with >3-year gaps in ACHD specialist care during their adulthood. Once we achieved thematic saturation [42] at UCSF, we recruited non-UCSF patients with convenience [43] and snowball sampling [44] to have representation from all regions of the US (Northeast, Southeast, Midwest, Southwest, and West). All patients underwent an informed consent process and completed a demographic questionnaire at the end of their interview. Clinicians included adult congenital cardiology and pediatric cardiology nurses, doctors, coordinators, board-certified patient advocates, and program leaders. Clinicians were recruited from UCSF and outside UCSF using a similar approach of snowballing, thematic saturation, and to have representation from all regions. Separate interview guides were developed for patients and clinicians using content domains within the COM-B framework and to explore opinions towards a digital tool and its key features (**Supplementary material**). Interviews were conducted over Zoom, each lasting 45-60 minutes, were audio-recorded and professionally transcribed.

#### Community-engaged approach

The goal of the research team in recruiting community advisory board (CAB) partners was to represent diverse perspectives in ACHD care. We primarily used convenience sampling to recruit patients from UCSF and snowball sampling to recruit patients outside UCSF, clinicians and advocacy representatives. We collaborated with the CAB partners to adapt the BCW components to identify ways to promote lifelong specialized care. CAB partners were involved in all aspects of intervention planning, design, and implementation, including reviewing findings from semi-structured interviews, drafting and finalizing tool features and branding, sharing personal experiences, and partnering throughout iterative prototype testing. They also assisted with recruitment of interview participants and contributed to implementation, evaluation, and dissemination activities. One patient partner (JV) led the creation of a design guide to ensure consistency and uniformity in materials, built the study website, and developed strategies to elevate participant experience. CAB members shared their stories as part of the intervention content and partnered as co-authors in scientific and community writing and dissemination. CAB members provided ongoing input through monthly hour-long meetings. Some CAB members also participated as test users in the pilot testing of the tool for iterative refinement. Meeting discussions were documented in study logs and analyzed by the research team to identify key themes and design recommendations. Community feedback was integrated iteratively with interview data to inform intervention components, branding, and content strategy.

### Data Analysis

We conducted thematic analysis of all semi-structured interviews. Each transcript was coded independently by at least two coders to identify themes and subthemes related to barriers and facilitators to lifelong specialized care and towards key aspects of digital tool. The two versions of coded transcripts were then compared to resolve discrepancies between coders and to generate a single, final coded transcript for each interview. We created a structured data matrix using Rapid Qualitative Analysis [45], an action-oriented method for qualitative data analysis conducted in Microsoft Excel, to efficiently produce results for real-world interventions. Using the matrix, cross-participant comparisons were made to identify determinants of ACHD specialized care and identify factors associated with use of the digital tool. We summarized the matrix findings via iterative group meetings of five team members (AA, KM, KB, JM, PA), which were then reviewed by the CAB members. Through ongoing, iterative review of CAB meeting discussions documented in study logs, we informed, planned, and refined the intervention components concurrent with their development.

### Ethics Approval

Ethical approval for this study was obtained from The University of California San Francisco Institutional Review Board (IRB # 22-36667). Verbal informed consent was obtained from interview participants for anonymized patient information to be published in this article. The National Clinical Trial number for this study is NCT06581484.

## Results

### Study population

We interviewed 54 participants (37 patients and 17 clinicians) to understand the determinants to lifelong ACHD specialized care and key features of the digital tool (**Table 3**).

**Table 3:** Characteristics of Study Participants.

| **Characteristic** | **Patients (N=37)** | **Clinicians (N=17)** |
| --- | --- | --- |
| Age, median (range) | 32 (18-65) | NA |
| Female, n (%) | 21 (57%) | - |
| Race/Ethnicity, n (%) |  |  |
| - Asian | 8 (21%) | - |
| - Black/ African American | 6 (15%) | - |
| - Hispanic/ Latino | 7 (18%) | - |
| - White | 13 (35%) | - |
| - Other/Multiple | 3 (8%) | - |
| Education ≤ some college, n (%) | 14 (41%) | - |
| Care gap ≥ 3 years, n (%) | 13 (35%) | - |
| Recruitment Source |  |  |
| - UCSF | 27 (73%) | 9 (52%) |
| - Community/snowball | 10 (27%) | 8 (48%) |
| Clinician Roles |  |  |
| - ACHD/Pediatric Cardiologists | - | 8 (47%) |
| - Nurses/NPs | - | 6 (35%) |
| - Other (coordinators, advocates) | - | 3 (18%) |

The CAB included 21 members comprising 11 patients or family members who are racially diverse and from all regions of the US, six pediatric or ACHD physicians, a social worker, an ACHD nurse, two nurse practitioners, and six advocacy representatives, some of whom represent more than one role. Advocacy organizations included the Adult Congenital Heart Association (ACHA), Conquering CHD, The Mended Hearts, and Team Uncle Joe [46–49].

### CHD-related unique challenges to inform ACHD digital intervention design and features

The qualitative interviews and our CAB partners identified challenges uniquely faced by ACHD patients. These challenges centered around users’ perspectives, loneliness, feasibility, accessibly, credibility, and scalability (**Table 4**). They then helped identify design objectives and intervention features that could addresses these challenges. The interview participants also identified four patient-centered goals to consider during the design of the digital tool (**eTable 1**): i) Easy access to credible resources (How can we make it easier to connect patients with pre-existing resources); ii) Uplifting of patient voices (How can we uplift patient voices to create a personal connection); iii) Customization to patient needs (How can we cater to different needs within the community); and iv) Centering positivity and joy (How can we bring positivity and joy into our tool).

**Table 4.**
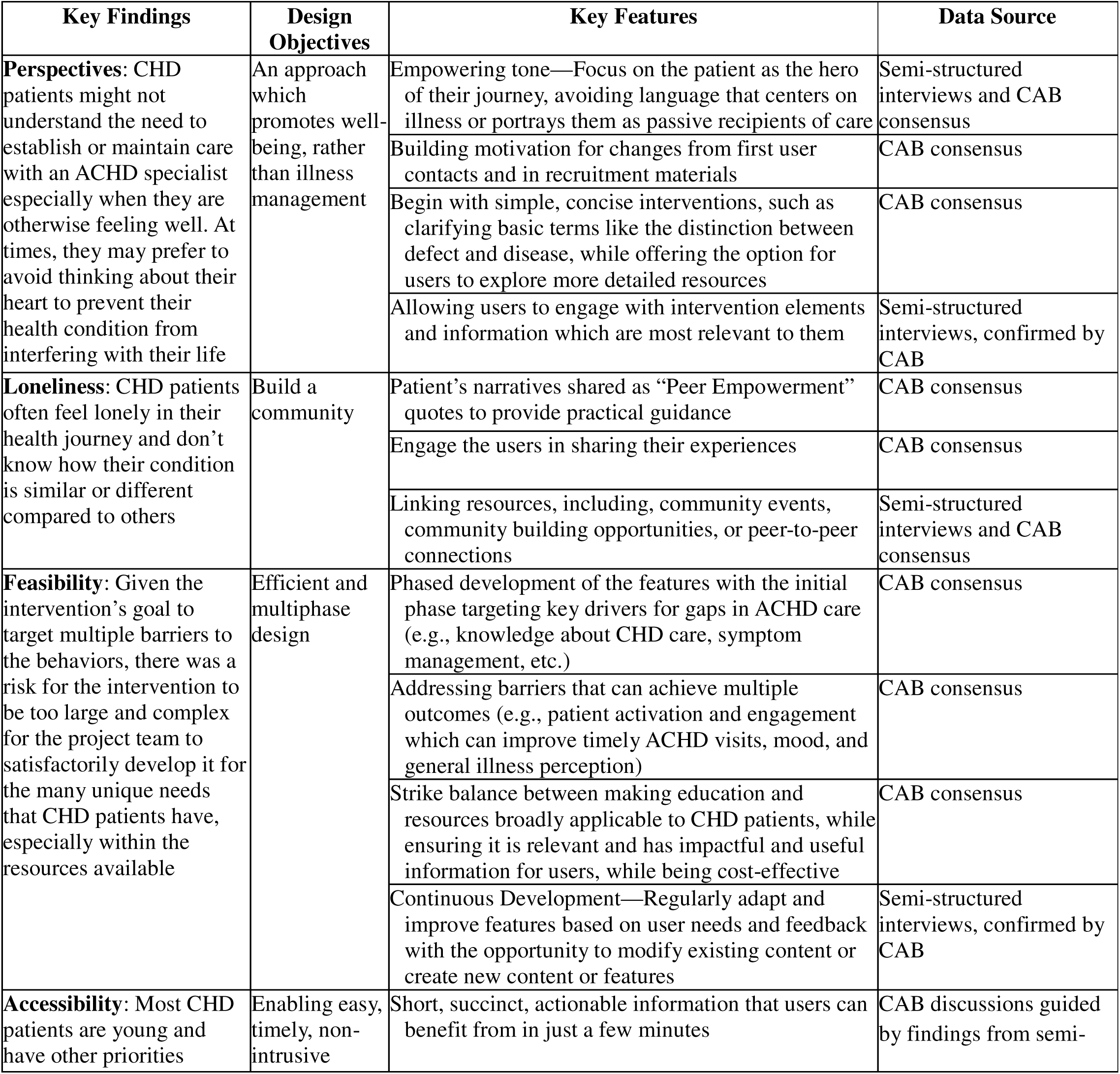
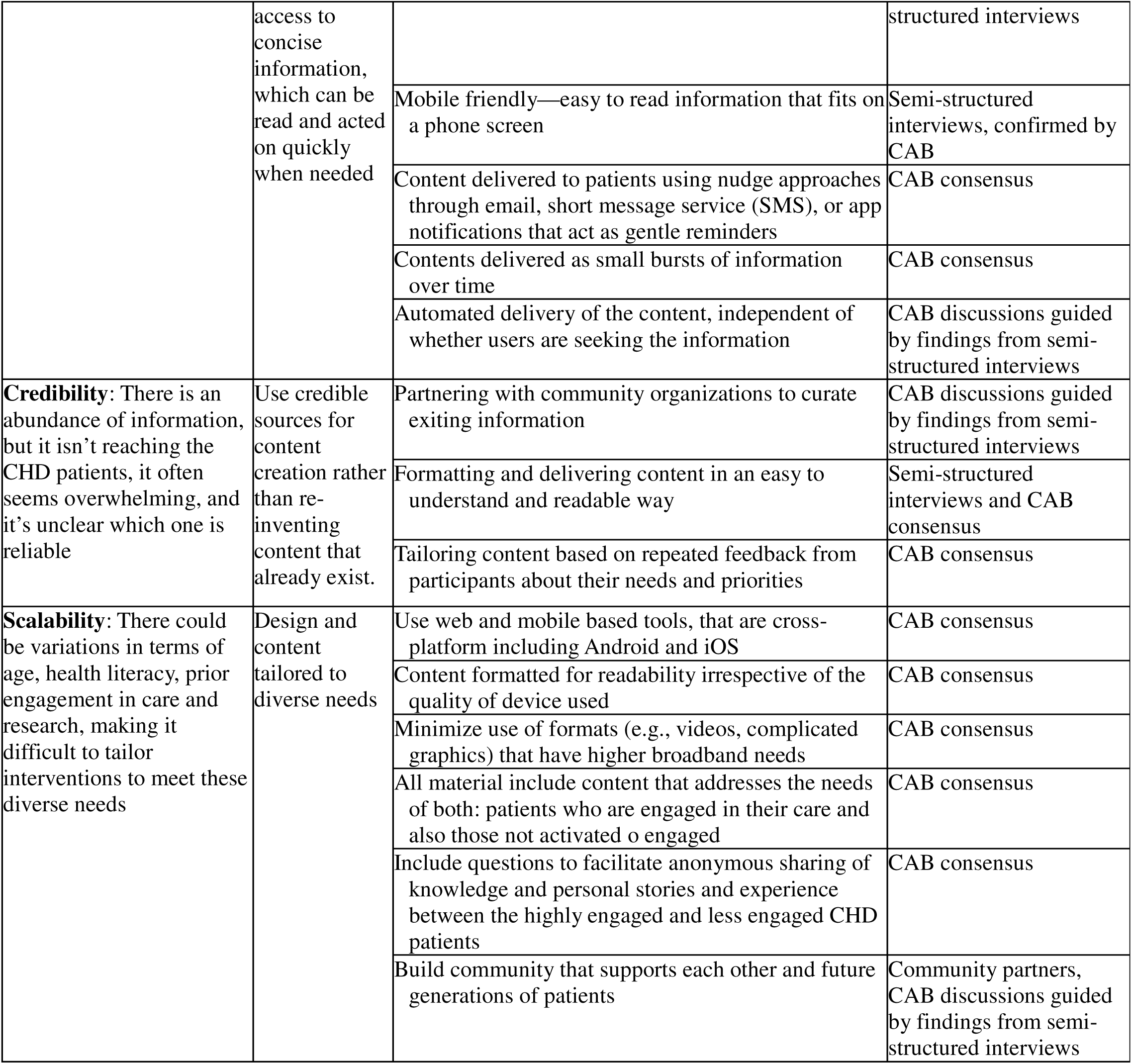
Digital Tool Design Objectives and Features for ACHD Specialized Care.

### Theory-driven intervention components

Using the BCW framework, we mapped barriers to specialized care (identified through interviews and CAB discussions) to behavioral targets and intervention functions (**Figure 2**). This systematic approach enabled us to select six BCW functions (Education, Training, Environmental Restructuring, Modeling, Persuasion, and Enablement) and translate them into specific intervention components. For example, educational modules with analogies explaining CHD conditions address capability barriers (lack of knowledge), while peer narratives simultaneously address social opportunity barriers (modeling self-advocacy) and motivation barriers (building confidence and hope). We further mapped these intervention components to intervention objectives to develop a theoretical framework showing how intervention functions are hypothesized to impact outcomes (**Figure 3**).

**Figure 2:**
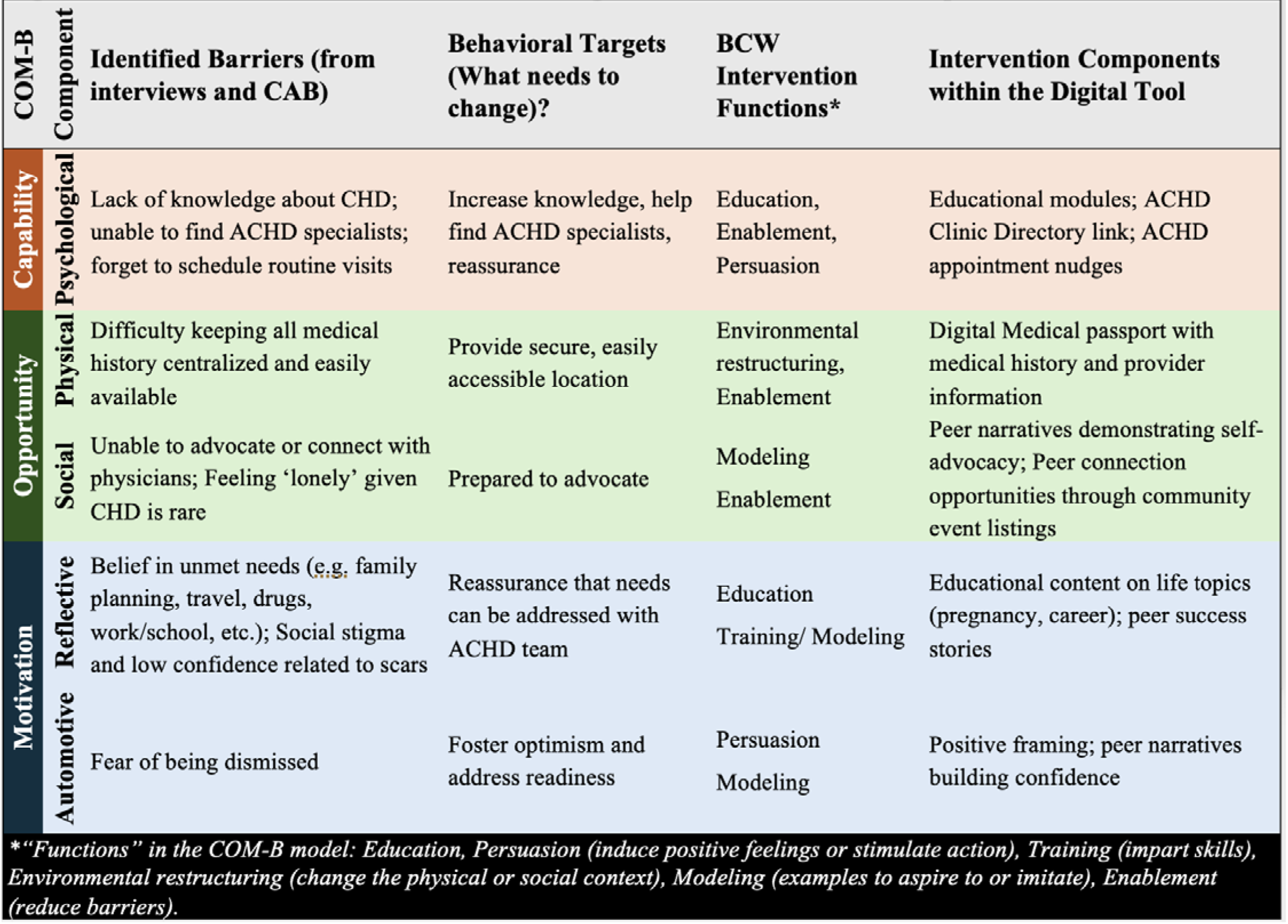
COM-B and BCW Framework for the Digital Intervention to Promote Specialized ACHD Care. Semi-structured interviews and community partners identified barriers to specialized ACHD care amenable to digital intervention and patient-centered goals. These were mapped to BCW to create a theory-driven approach for intervention development. This framework identified COM-B barriers to specialized care, behavioral targets to address those barriers, and intervention functions that informed the selection of specific intervention components within the digital tool.

**Figure 3:**
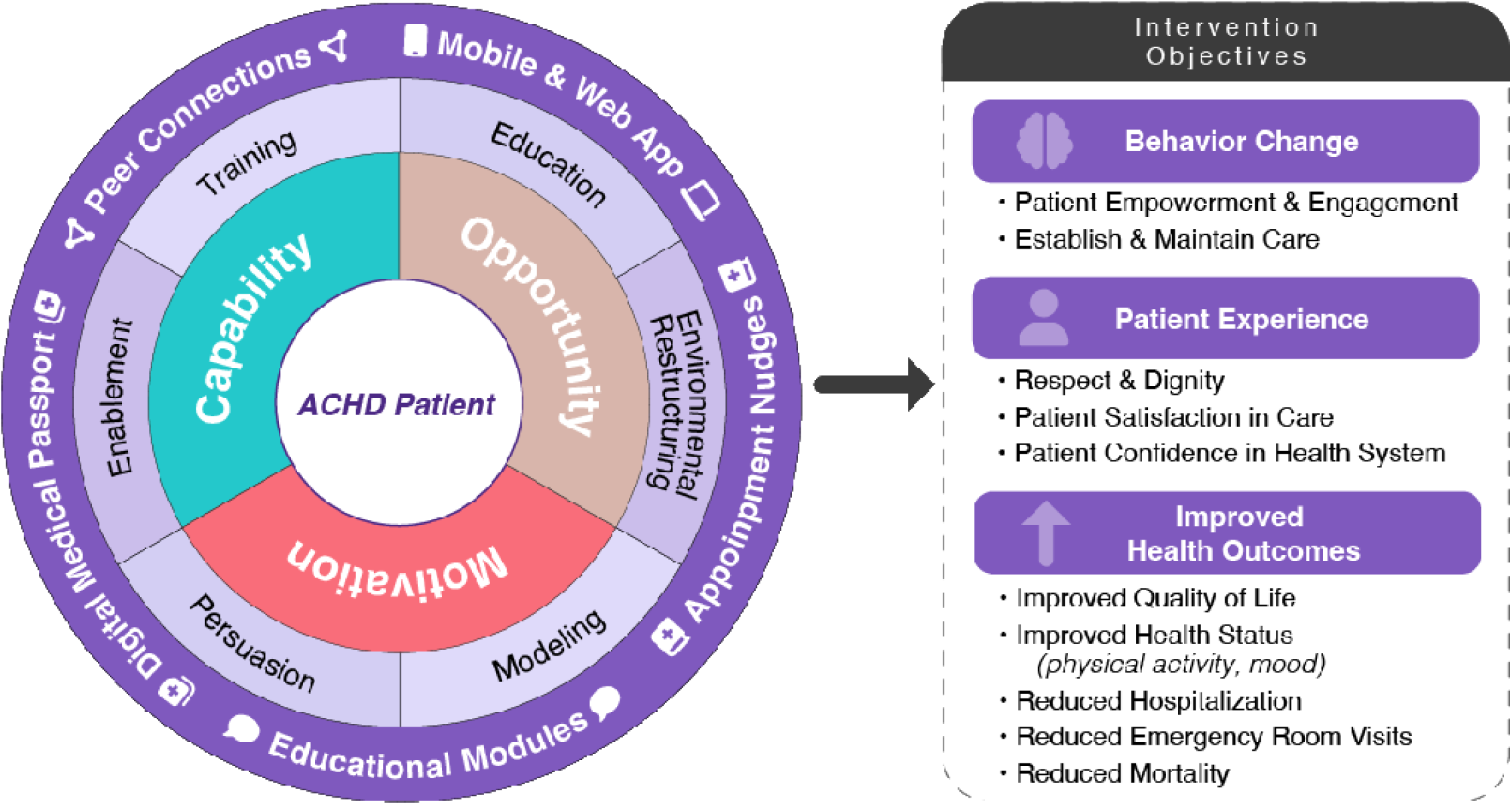
Theoretical framework Hypothesizing the Impact of Digital Health Intervention Components on Outcomes. The intervention components (educational modules, digital medical passport, peer connections, and appointment nudges), delivered via web and mobile app, function as tools for education, training, enablement, persuasion, modeling, and environmental restructuring to enhance the Capability, Opportunity, and Motivation (COM-B) domains of adult congenital heart disease (ACHD) patients. We hypothesize that these components will support patient activation and engagement skills and lifelong specialized ACHD care, ultimately improving patient experience and health outcomes.

### Design of the digital tool

#### Digital Research Platform

The digital tool was designed and hosted on the Eureka Research Platform (University of California, San Francisco), a HIPAA-secure, NIH-supported infrastructure that integrates data collection and intervention delivery within a single platform [50]. Participants enroll remotely via electronic consent using QR codes or web links (UCSF IRB#22-36667).

#### Branding

The community partners discussed various options for name and logo of the digital tool and narrowed them down to four potential names: My Congenital Heart Care, Empower My Congenital Heart, My Congenital Heart Guide, Uplift My Congenital Heart. The CAB members suggested to include an arrow in the logo to reflect the primary goal of uplifting and empowering patients. Red, blue, and purple colors were chosen to reflect the diversity within CHD lesions and their uniqueness from other heart diseases. The design team developed four branding prototypes reflecting CAB priorities: upward arrows symbolizing empowerment and red-blue-purple color schemes representing CHD diversity (**eFigure 1**). Through CAB voting, ’Empower My Congenital Heart (EMCH)’ with an integrated heart-and-arrow logo (option B-2) was selected.

#### Participant flow within EMCH

The intervention design enables recruitment through email, clinic flyers, social media, or community outreach using study-specific QR codes. After registration and electronic consent, participants engage via smartphone or web browser. Activities are delivered every 2 months, a schedule selected by the CAB to balance engagement and participant burden. Each activity cycle remains available for 2 months and includes surveys, educational modules, and optional linkages to wearables, smartphone data, and patient portals (**Figure 4**). Surveys gather self-reported data, feedback, and allow them to share stories or questions (**eTable 2**). Modules provide concise guidance from ACHD experts and patients to enhance confidence and knowledge in navigating CHD care (see below). Optional linkages enable participants to share their health data, supporting research and understanding of ACHD patient experiences and behaviors. For example, during the first 2-month cycle (enrollment to month 2), participants complete baseline demographic surveys, after which Module 1 (A Guide to Choosing Your Care Team) and optional linkages become accessible. During months 2-4, quality-of-life surveys become available, followed by Module 2 (Navigating Cardiologist Appointments) and optional linkages, if not already completed.

**Figure 4:**
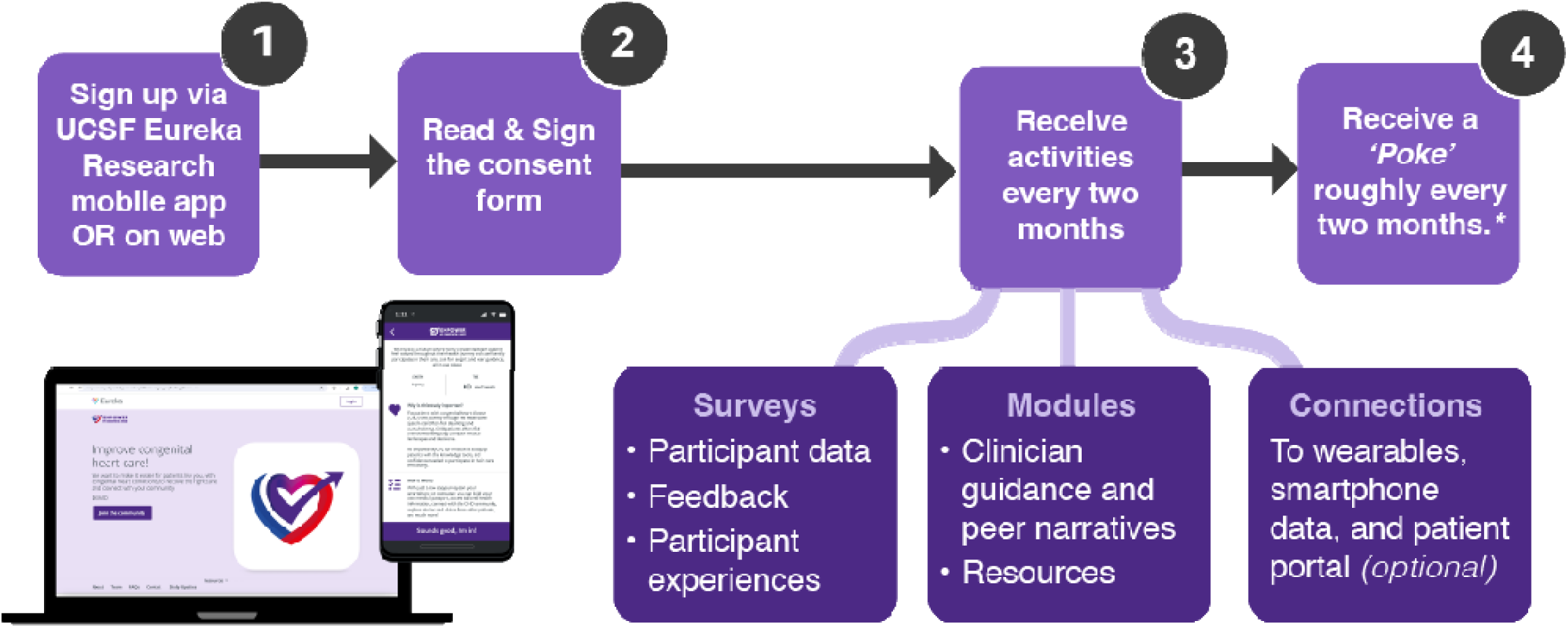
Participant Flow Through EMCH. EMCH is accessible via the UCSF Eureka Research mobile app or web platform. Participants first create a UCSF Eureka Research account (Step 1), then review and electronically sign the EMCH consent form (Step 2). Following consent, participants receive study activities every 2 months (Step 3), including surveys (participant data, feedback, and participant experiences), modules (clinician guidance and peer narratives, resources on topics related to CHD and health system navigation), and optional connections to wearables, smartphone data, and patient portals. Participants also receive a “Poke” notification roughly every 2 months to remind them of new activities (Step 4). Representative screenshots show the web-based landing page and mobile-app study information page.

#### Core intervention components within EMCH (Figure 5)

The following sections describe how each intervention component operationalizes the BCW intervention functions identified in **Figure 3**.

1. <u>Educational modules</u> (addressing **capability** and **motivation** barriers): Co-designed with the CAB, modules are developed around three core takeaways aligned with the COM-B framework: increasing knowledge about CHD and navigating health system (capability), enabling access to ACHD specialists (opportunity), and incorporating peer narratives to build motivation. Modules combine concise educational content with clinician-derived “Empowerment” messages and patient-derived “Peer Empowerment” stories to reinforce active self-management through confidence, social influence, and optimism, and may include brief interactive elements (e.g., trivia). Examples of “Empowerment” quote include “Often, over time you build strong relationships with the nurses, social workers, and care coordinators in the team who can help with insurance approvals, disability forms, etc.” while that of Peer Empowerment include “When deciding whether to go to the emergency room, I call my cardiologist while on the way to discuss the situation. If my cardiologist says I don’t need to go, I simply turn around. This way, I’m already en route if I do need to be seen.” (Figure 5). Community partners identified seven priority themes to support health system navigation based on user needs that determined the content of the modules (**eTable 3).** The first seven modules (Year 1) cover core components related to CHD education and health-system navigation, mapped to BCW constructs. After completing the year 1 core modules, participants may continue to engage with additional modules (such as psychosocial concerns, work or travel considerations, diet, exercise, pregnancy/family planning, dental care, etc.); brief bimonthly “pokes” featuring peer stories, study updates, or CHD research opportunities; appointment nudges; or through surveys where they can share their stories. All materials are designed using a design guide (**eTable 4**) to ensure uniformity and consistency and enhance readability. Currently, all participants receive the same core educational modules in the same sequence. However, “Knowledge About My CHD Condition” module allows self-directed personalization where participants can select their specific CHD diagnosis (e.g., atrial septal defect, tetralogy of Fallot, single ventricle, etc.) and engage only with the content relevant to them.
2. <u>Appointment planning nudges</u> (addressing **capability** barriers of forgetting routine visits): Embedded within the annual surveys, the gentle nudges prompt scheduling or completion of ACHD specialist visits.
3. <u>Personal digital medical passport</u> (addressing **physical opportunity** barrier of centralized information): The passport is automatically generated from baseline self-reported data about congenital heart diagnosis, pacemaker/defibrillator device type and manufacturer (if applicable), other medical comorbidity information, ACHD cardiologist name and clinic location, primary care physician name and location, and any additional relevant health details (e.g. oxygen saturation, allergies, etc.) (**Figure 6**). The passport serves as a centralized hub for all medical and provider information while also linking participants to curated community events, all EMCH resources (modules), the ACHD provider directory, and a secure feedback and contact form. The passport is always accessible via smartphone within the EMCH app and can be shared with clinicians by displaying on-screen. As the passport data is self-reported and build from baseline surveys, users are encouraged to verify accuracy with their ACHD team while completing these surveys (functionality to update the passport information is under development).
4. <u>Peer connections and community support</u> (addressing **social opportunity and motivation** barriers): Peer Empowerment quotes integrated throughout modules provide modeling for self-advocacy and build confidence. Community event listings connect participants to other ACHD patients by providing information about the local CHD walks, camps, support groups, and social media channels to reduce isolation. Before final inclusion, all peer-contributed content is thoroughly reviewed by the CAB members and the research team for authenticity, relevance, medical accuracy, tone, alignment with design objectives, privacy protection, and for removal of any distressing content. Currently, EMCH does not include open peer-to-peer messaging or unmoderated forums to ensure content quality and participant safety. All peer content is curated and vetted before being shared with the participants.

**Figure 5:**
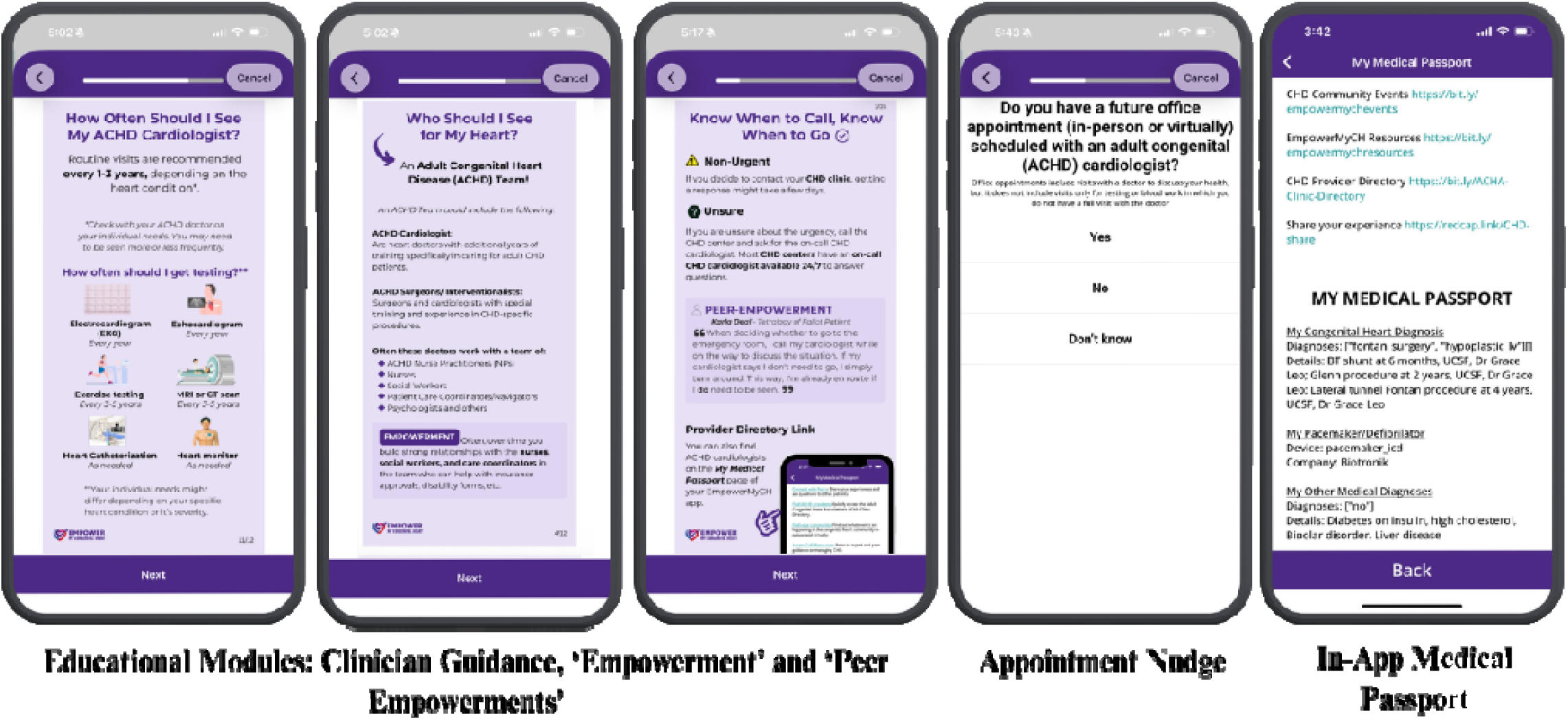
Representative Core Intervention Components within EMCH. The screenshots show representative screens of the educational modules (with clinician guidance, “Empowerment”, and patient-derived “Peer Empowerment” stories), appointment planning nudges (embedded within the surveys), and personal digital medical passport (a centralized hub for all medical and provider information while also linking participants to curated community events, all EMCH modules, the ACHD provider directory, and a secure feedback and contact form). Peer connections component of EMCH is integrated within the modules and community event listings.

**Figure 6:**
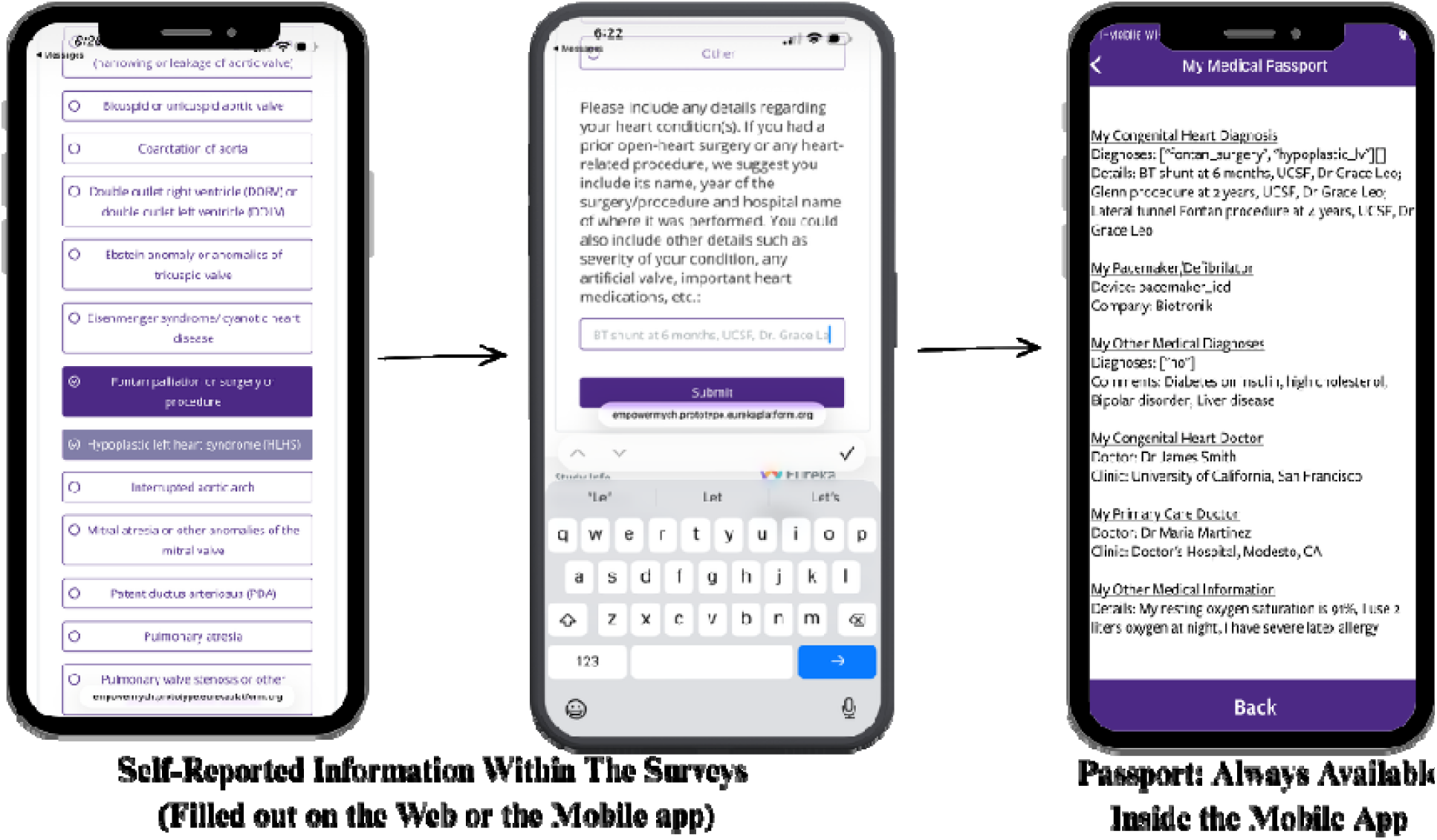
Digital Medical Passport Built Using Self-Reported Data. This figure illustrates the type of health information easily accessible to participants at any time through the EMCH mobile app. By completing a series of surveys with self-reported data, participants build their own digital medical passport, a personalized hub for their vital health and provider information.

#### Iterative Refinements During Intervention Development

Throughout development, intervention features were refined based on pilot testing, CAB feedback, and adherence to digital health equity principles (**Table 5**). These refinements demonstrate the responsive, user-centered development process that shaped the final EMCH design.

**Table 5:** Iterative Refinements During Intervention Development.

| **Table 5: Iterative Refinements During Intervention Development** |  |  |  |
| --- | --- | --- | --- |
| **Initial Design** | **Refinement** | **Rationale** | **Data Source** |
| Monthly delivery of the module | Modules delivered every 2 months | Reduce participant burden while maintaining engagement | CAB feedback |
| Colorful icons/ imagery | Flat, minimalist icons | Enhance readability, reduce bandwidth needs, improve accessibility | Digital health equity principles + CAB feedback |
| Frequent reminders (9 total: 3 email, 3 push, 3 texts per module) | Reduced to 3 total reminders per module | Minimize intrusiveness, respect participant time | User testing + CAB feedback |
| Reminders sent until all activities completed (survey and modules) | Reminders sent only if module incomplete; most surveys are not re-prompted | Minimize intrusiveness for data collection while providing opportunities to engage with modules | CAB feedback |
| Digital passport surveys delivered at 3 months post enrollment | Digital passport surveys delivered at the time of enrollment | Maximize early engagement with digital passport from first access | User testing + CAB feedback |
| Clinician guidance and peer narratives labeled as “Tips” | Clinician narratives as “Empowerment” quotes; peer narratives as “Peer Empowerment” quotes | Emphasize empowerment and positivity as primary intervention goals | CAB feedback |
| Module content primarily informational | Added interactive elements (trivia questions, action items) within the modules | Enhance engagement and encourage clear, actionable takeaways | CAB feedback |
| Digital passport to include comprehensive data (CHD history, other medical history, all providers, medications, insurance) | Digital passport limited to pertinent medical and provider information | Simplify content to enhance usability and relevance; reduce data entry burden | Eureka platform team + CAB feedback |

#### Engagement in EMCH

Engagement is supported through multi-modal communication (app notifications, email, SMS), with up to three reminders per activity cycle sent at varying times to accommodate different schedules and time zones. A unique feature is the bi-monthly ’Poke’: a brief, variable-timing communication featuring peer stories, study updates, CHD research opportunities, or community-shared experiences. Content variability and unpredictable timing of the Pokes was a feature adapted from established Eureka workflow that enable intermittent reinforcement and maintain interest [50].

### Data Security and Privacy

The Eureka Platform uses a 128-bit secure socket layer (SSL) protocol that protects all data transmission between the app and server users. Data is stored on HIPAA-compliant servers managed by UCSF with restricted access to essential research personnel, using multi-factor authentication with role-based permissions. Survey responses and engagement metrics are stored as separate csv files using a unique identifier but no personal identifiable information. Only authorized research team can link the de-identified data to individual participants. Participants can withdraw from the study at any time through the app or by contacting the research team. Upon withdrawal, participants can request deletion of their data or allow de-identified data to remain for research purposes. Participants control whether their stories can be shared as Peer Empowerment content (explicit opt-in required). Wearable device data (FitBit) and patient portal connections require separate participant authorization and are governed by those platforms’ privacy policies in addition to EMCH protections. Study data will be retained per UCSF IRB and NIH data sharing policies, after which it will be securely destroyed or permanently de-identified.

## Discussion

### Principal Results

We describe a theory-based, community co-designed approach to developing EMCH, a digital intervention promoting confidence and skills to enhance engagement in lifelong ACHD specialized care. To our knowledge, this is the first cross-platform (Android and iOS) digital tool specifically designed to address ACHD-specific barriers to lifelong specialized care. Our approach integrated evidence-based behavioral frameworks (COM-B/BCW) with extensive qualitative research (n=54) and community-based participatory design to map barriers to engagement and develop targeted intervention components. The CBPR approach ensured patient and clinician perspectives shaped all design decisions. EMCH enables convenient, passive content access using user-friendly design and behavioral economics (nudge) principles while fostering peer connections and community contribution.

### Clinical Relevance and Expected Impact

Existing CHD digital interventions focus narrowly on symptom monitoring, exercise support, or transition readiness in young adults and lack theoretical grounding or rigorous evaluation [51,52]. EMCH addresses the full adult lifespan (18+ years) and targets the complete spectrum of barriers to ongoing specialist care, including those who never transitioned from pediatric care or were lost to follow-up. The intervention’s systematic foundation in behavioral theory (COM-B/BCW) distinguishes it from information-only approaches by addressing psychological capability, social opportunity, and motivation simultaneously. If effective, EMCH could promote lifelong specialized care and reduce preventable ACHD complications, emergent hospitalizations, and mortality associated with care gaps.

### Scalability and Sustainability

We aim to recruit and engage a representative ACHD population, particularly those with a high risk for gaps in care (young adults, men, rural residence, lower socioeconomic status, those living far away from CHD centers, etc.) and those not previously engaged in research. Mobile phones are nearly ubiquitous - 96% of Americans (and 94% of ACHD patients) in their twenties and thirties own a mobile phone, and reliance on them for online access is especially common among young adults, non-Whites, and lower-income Americans [22,53,54]. This provides us with a huge opportunity to enhance the reach and scalability of our digital health intervention to various subgroup of ACHD patients. With the use of digital health equity and diffusion of innovation frameworks and the versatility of options for our digital intervention (cross-platform low bandwidth content delivery with opportunities for translation, etc.), our intervention is likely to be relevant to the most representative population. Digital tools offer scalable solutions to supplement clinic-based care. EMCH was designed to augment, not replace, individualized patient-provider interactions by building engagement skills outside clinic visits. We selected the Eureka Digital Research Platform for its sustainability (NIH-supported, UCSF-owned), integration of recruitment through intervention delivery, and cost-efficiency supporting hundreds of concurrent studies. Unlike clinic-based navigator programs requiring ongoing personnel costs, EMCH’s automated content delivery and peer-generated stories provide sustainable engagement with minimal marginal cost per additional user. This infrastructure enables cost-efficient delivery at scale while maintaining intervention fidelity.

### How EMCH Differs from Existing Approaches

We selected BCW over alternative frameworks (e.g., Intervention mapping or the Consolidated Framework for Implementation Research) [55,56], because it provides flexible, evidence-based guidance for digital behavior change interventions without being overly prescriptive [57,58]. The BCW process enabled transparent documentation of our intervention’s theory of action by systematically mapping behavioral barriers to specific intervention components using shared taxonomic language [30]. Integrating community-engaged approach with behavioral theory proved essential for moving beyond merely identifying known barriers [28]. Qualitative methods and community engagement revealed not just what barriers exist, but how to address them effectively. For example, while we knew many ACHD patients lack awareness of specialist care needs [5], interviews and CAB feedback showed that evidence-based information alone was insufficient. Participants needed peer stories and real-life scenarios to internalize the message’s relevance. This insight led to “Empowerment” and “Peer Empowerment” quotes becoming core intervention components, using modeling to build confidence and demonstrate care’s importance when patients feel well. The combined theory- and community-engaged approach can be resource-intensive but adaptable. Teams with limited resources can supplement qualitative work with rapid stakeholder consultation while maintaining the framework’s core strengths: preventing reinvention of existing content, simultaneously addressing engagement and implementation barriers, and ensuring intervention components meet users’ actual needs.

### Limitations

EMCH addresses patient-level barriers but cannot overcome system-level challenges such as ACHD specialist shortages, insurance coverage gaps, or geographic distance from specialized centers. The intervention is likely most appropriate for stable patients capable of self-management with remote guidance; patients with complex acute needs or severe cognitive impairment require in-person clinical support that digital tools cannot replace. The digital tool requires a smartphone or computer with internet access. While limited digital literacy is uncommon in the relatively young ACHD population (94% own smartphones) [22], and EMCH is available on web and native iOS/Android platforms with low-bandwidth design, the most vulnerable patients with care gaps may face barriers beyond digital intervention scope. However, by reducing routine educational burden for digitally engaged patients, EMCH may enable clinical teams to prioritize in-person resources for those with greater needs. The digital passport currently captures baseline data only and does not allow users to update information as their medical status changes, which may limit accuracy over time. We are developing update functionality for future versions and exploring integration with electronic health records to auto-populate verified clinical data. Although we incorporated diverse perspectives during development, we may not have captured all viewpoints from populations or organizations not initially included. The EMCH infrastructure supports continuous adaptation through participant feedback and research collaborations, enabling rapid implementation of new components (e.g., mental health modules, culturally adapted content). Currently, individuals with intellectual disabilities are excluded due to unique accommodation needs requiring caregiver-focused approaches.

### Next Steps

EMCH launched in September 2024 and has enrolled 500+ participants. Ongoing process evaluation is assessing acceptability, usability, and preliminary engagement patterns across diverse ACHD populations. We are developing Spanish-language adaptation to reach the 15-20% of ACHD patients who are primarily Spanish-speaking. Following process evaluation, we will conduct a randomized controlled trial evaluating EMCH’s effectiveness on completing ACHD specialist visits and improving knowledge, self-efficacy, and social connectedness. If proven effective, EMCH will be made available to ACHD patients nationally through partnerships with our community advocacy organizations and in diverse clinical settings.

## Conclusions

The theory-based, community co-designed EMCH digital tool provides ACHD patients with evidence-based resources, peer support, and practical skills to become activated and engaged partners in their care. Ongoing process evaluation will identify the acceptability, feasibility, and satisfaction with the EMCH intervention components, enabling refinement and tailoring for future hybrid effectiveness trials. If proven effective, EMCH’s approach can be adapted for other chronic conditions requiring lifelong specialized care, as the behavioral frameworks and intervention strategies address universal barriers to care engagement. The EMCH infrastructure also enables rapid recruitment of diverse ACHD populations for future research and health policy initiatives.

## Data Availability

All data produced in the present study are available upon reasonable request to the authors.

## Acknowledgments

The authors would like to acknowledge the important contributions of the patients, families, health care professionals, and advocacy representatives who partnered as our community advisory board.

## Disclosure of Delegation to Generative AI

The authors declare the use of generative AI in the research and writing process. According to the GAIDeT taxonomy (2025), the following tasks were delegated to GAI tools under full human supervision:

- Proofreading and editing

- Summarizing text

- Reformatting

The GAI tool used was: Claude Sonnet 4.5.

Responsibility for the final manuscript lies entirely with the authors.

GAI tools are not listed as authors and do not bear responsibility for the final outcomes.

Declaration submitted by: Anushree Agarwal

## Author Contributions

A.A. and J.V. contributed to all aspects of the study design, and in preparing the initial drafts and revisions of the paper.

K.B contributed to the recruitment and retention of study participants, qualitative analysis, development of study activities, and study regulatory management.

K.M. contributed to the qualitative interview process, identification of necessary design objectives and intervention features for EMCH.

J.M, K.R., K.M., and P.A. contributed to qualitative analysis, conduct of the study and critical review of the manuscript.

P.K. contributed to the technical development and adaptation of EMCH study activities on the UCSF Eureka Research platform.

K.L., K.P., M.D.N, K.B.J., L.R., P.M., M.O, G.M.M, and M.G. contributed to the overall study design and critical review of the manuscript.

Supplementary Information is available for this paper.

Correspondence and requests for materials should be addressed to.

## Funding

Research reported in this manuscript was supported by the National Heart, Lung, and Blood Institute of the National Institutes of Health under Award Number K23HL151866 (Agarwal). The content is solely the responsibility of the presenters and does not necessarily represent the official views of the National Institutes of Health. Greg Marcus discloses support for the Eureka Research Platform from National Institute of Biomedical Imaging and Bioengineering under Award Number 3U2CEB021881-05S1.

## Competing Interests

Dr. Mark Norris is a Consultant for American College of Cardiology since 2024.

## Abbreviations

ACHD: Adult Congenital Heart Disease
BCW: Behavior Change Wheel theoretical model
CAB: Community Advisory Board
COM-B: Capability, Opportunity, Motivation for Behavior Change
CHD: Congenital Heart Disease
eConsent: electronic consent
EMCH: Empower My Congenital Heart
HIPAA: Health Insurance Portability and Accountability Act
iOS: iPhone Operating System
NIH: National Institutes of Health
SMS: Short Message Service
QR code: Quick response code
UCSF: University of California, San Francisco
US: United States

## Notes

### Author Declarations

Ethics/IRB of University of California, San Francisco gave ethical approval for this work.

### Summary of Updates

Figures were updated and the clarity of the paper was improved.

## References

1. Gilboa SM, Devine OJ, Kucik JE, et al. Congenital Heart Defects in the United States: Estimating the Magnitude of the Affected Population in 2010. Circulation. 2016;134(2):101–109. doi:10.1161/CIRCULATIONAHA.115.019307

2. Khairy P, Ionescu-Ittu R, Mackie AS, Abrahamowicz M, Pilote L, Marelli AJ. Changing mortality in congenital heart disease. Journal of the American College of Cardiology. 2010;56(14):1149–1157.

3. Stout KK, Daniels CJ, Aboulhosn JA, et al. 2018 AHA/ACC Guideline for the management of adults with congenital heart disease: executive summary: a report of the American College of Cardiology/American heart association task force on clinical practice guidelines. Circulation. Published online 2018:CIR. 0000000000000602.

4. Gurvitz M, Krieger EV, Fuller S, et al. 2025 ACC/AHA/HRS/ISACHD/SCAI Guideline for the Management of Adults With Congenital Heart Disease: A Report of the American College of Cardiology/American Heart Association Joint Committee on Clinical Practice Guidelines. Circulation. Published online December 18, 2025:CIR.0000000000001402. doi:10.1161/CIR.0000000000001402

5. Gurvitz M, Valente AM, Broberg C, et al. Prevalence and predictors of gaps in care among adult congenital heart disease patients: HEART-ACHD (The Health, Education, and Access Research Trial). Journal of the American College of Cardiology. 2013;61(21):2180–2184.

6. Khan AM, McGrath LB, Ramsey K, Agarwal A, Slatore CG, Broberg CS. Distance to Care, Rural Dwelling Status, and Patterns of Care Utilization in Adult Congenital Heart Disease. Pediatric cardiology. Published online 2021:1–9.

7. Mackie AS, Ionescu-Ittu R, Therrien J, Pilote L, Abrahamowicz M, Marelli AJ. Children and adults with congenital heart disease lost to follow-up: who and when? Circulation. 2009;120(4):302–309. doi:10.1161/CIRCULATIONAHA.108.839464

8. Bayne J, Duan R, Rudov L, et al. Understanding gaps in guideline-recommended adult congenital heart disease care: Data from 12 US health care centers. Am Heart J. 2025;291:53–62. doi:10.1016/j.ahj.2025.08.002

9. Yeung E, Kay J, Roosevelt GE, Brandon M, Yetman AT. Lapse of care as a predictor for morbidity in adults with congenital heart disease. International journal of cardiology. 2008;125(1):62–65.

10. Mylotte D, Pilote L, Ionescu-Ittu R, et al. Specialized adult congenital heart disease care: the impact of policy on mortality. Circulation. 2014;129(18):1804–1812. doi:10.1161/CIRCULATIONAHA.113.005817

11. Agarwal A, Duan R, Bayne J, et al. Impact of Adult Congenital Heart Disease Specialist Visits on Emergent Admissions: Evidence for Guidelines. JACC Adv. 2025;4(8):102021. doi:10.1016/j.jacadv.2025.102021

12. Agarwal A, Macholl K, Qian A, et al. Patient- and Clinician-Solutions to Improve Specialized ACHD Care: A Theory-Based Approach. Preprint posted online April 3, 2025. doi:10.1101/2025.04.01.25325065

13. John AS, Jackson JL, Moons P, et al. Advances in managing transition to adulthood for adolescents with congenital heart disease: a practical approach to transition program design: a scientific statement from the American Heart Association. Journal of the American Heart Association. 2022;11(7):e025278.

14. Bratt EL, Mora MA, Sparud-Lundin C, et al. Effectiveness of the STEPSTONES Transition Program for Adolescents With Congenital Heart Disease—A Randomized Controlled Trial. Journal of Adolescent Health. Published online 2023.

15. Gaydos SS, Chowdhury SM, Judd RN, McHugh KE. A transition clinic intervention to improve follow-up rates in adolescents and young adults with congenital heart disease. Cardiology in the young. 2020;30(5):633–640.

16. Mackie AS, Rempel GR, Kovacs AH, et al. Transition intervention for adolescents with congenital heart disease. Journal of the American College of Cardiology. 2018;71(16):1768–1777.

17. Thomet C, Schwerzmann M, Budts W, et al. Transfer and transition practices in 96 European adult congenital heart disease centres. International journal of cardiology. 2021;328:89–95.

18. Fernandes SM, Khairy P, Fishman L, et al. Referral patterns and perceived barriers to adult congenital heart disease care: results of a survey of US pediatric cardiologists. Journal of the American College of Cardiology. 2012;60(23):2411–2418.

19. Fernandes SM, Verstappen A, Clair M, et al. Knowledge of Life-Long Cardiac Care by Adolescents and Young Adults with Congenital Heart Disease. Pediatric cardiology. 2019;40(7):1439–1444.

20. Free C, Phillips G, Watson L, et al. The effectiveness of mobile-health technologies to improve health care service delivery processes: a systematic review and meta-analysis. PLoS Med. 2013;10(1):e1001363.

21. Low JK, Manias E. Use of technology-based tools to support adolescents and young adults with chronic disease: systematic review and meta-analysis. JMIR mHealth and uHealth. 2019;7(7):e12042.

22. Lopez KN, O’connor M, King J, et al. Improving transitions of care for young adults with congenital heart disease: mobile APP development using formative research. JMIR formative research. 2018;2(2):e16.

23. Heath S. Chronic disease management apps for teens need stakeholder input. Patient Satisfaction News. 2016. 2016;2023(September 27,). https://patientengagementhit.com/news/chronic-disease-management-apps-for-teens-need-stakeholder-input

24. Liang L, Bernhardsson S, Vernooij RW, et al. Use of theory to plan or evaluate guideline implementation among physicians: a scoping review. Implementation Science. 2017;12(1):26.

25. Handley MA, Abascal Miguel L, Thompson LM, Velloza J. Bridging community-engaged research and implementation science methods to advance public health practice. Bundesgesundheitsbl. 2025;68(7):797–808. doi:10.1007/s00103-025-04079-5

26. Riccardi M, Pettinicchio V, Di Pumpo M, et al. Community-based participatory research to engage disadvantaged communities: Levels of engagement reached and how to increase it. A systematic review. Health Policy. 2023;137:104905. doi:10.1016/j.healthpol.2023.104905

27. Baker TB, Gustafson DH, Shah D. How Can Research Keep Up With eHealth? Ten Strategies for Increasing the Timeliness and Usefulness of eHealth Research. J Med Internet Res. 2014;16(2):e36. doi:10.2196/jmir.2925

28. Yardley L, Morrison L, Bradbury K, Muller I. The person-based approach to intervention development: application to digital health-related behavior change interventions. J Med Internet Res. 2015;17(1):e30. doi:10.2196/jmir.4055

29. Cooper C, Watson K, Alvarado F, et al. Community Engagement in Implementation Science: the Impact of Community Engagement Activities in the DECIPHeR Alliance. Ethn Dis. 2023;DECIPHeR(Spec Issue):52–59. doi:10.18865/ed.DECIPHeR.52

30. Michie S, Stralen MMV, West R. The behaviour change wheel: a new method for characterising and designing behaviour change interventions. Implementation science. 2011;6(1):1–12.

31. Cane J, O’Connor D, Michie S. Validation of the theoretical domains framework for use in behaviour change and implementation research. Implement Sci. 2012;7:37. doi:10.1186/1748-5908-7-37

32. Richardson S, Lawrence K, Schoenthaler AM, Mann D. A framework for digital health equity. NPJ digital medicine. 2022;5(1):119.

33. Oldenburg B, Glanz K. Diffusion of innovations. Health Behavior and Health Education-Theory Research, and Practice. Published online 2008:313–330.

34. Liang SY, Stults CD, Jones VG, et al. Effects of Behavioral Economics–Based Messaging on Appointment Scheduling Through Patient Portals and Appointment Completion: Observational Study. JMIR Hum Factors. 2022;9(1):e34090. doi:10.2196/34090

35. Jackson C, Eliasson L, Barber N, Weinman J. Applying COM-B to medication adherence: a suggested framework for research and interventions. European Health Psychologist. 2014;16(1):7–17.

36. Alexander KE, Brijnath B, Mazza D. Barriers and enablers to delivery of the Healthy Kids Check: an analysis informed by the Theoretical Domains Framework and COM-B model. Implementation Science. 2014;9(1):1–14.

37. Whittal A, Störk S, Riegel B, Herber OR. Applying the COM-B behaviour model to overcome barriers to heart failure self-care: A practical application of a conceptual framework for the development of complex interventions (ACHIEVE study). European Journal of Cardiovascular Nursing. 2021;20(3):261–267.

38. McQuaid D, Doyle J. Designing digital behaviour change interventions to support older adults managing cardiac conditions. In: Proceedings of the 32nd International BCS Human Computer Interaction Conference 32. 2018:1–6.

39. Jatau AI, Peterson GM, Bereznicki L, et al. Applying the capability, opportunity, and motivation behaviour model (COM-B) to guide the development of interventions to improve early detection of atrial fibrillation. Clinical Medicine Insights: Cardiology. 2019;13:1179546819885134.

40. Sharry JM, Murphy PJ, Byrne M. Implementing international sexual counselling guidelines in hospital cardiac rehabilitation: development of the CHARMS intervention using the Behaviour Change Wheel. Implementation Science. 2016;11(1):1–11.

41. Higginbottom GMA. Sampling issues in qualitative research. Nurse Researcher (through 2013). 2004;12(1):7.

42. Saunders B, Sim J, Kingstone T, et al. Saturation in qualitative research: exploring its conceptualization and operationalization. Quality & quantity. 2018;52(4):1893–1907.

43. Golzar J, Noor S, Tajik O. Convenience Sampling. IJELS. 2022;1(2). doi:10.22034/ijels.2022.162981

44. Snowball Sampling. In: The SAGE Dictionary of Social Research Methods. SAGE Publications, Ltd; 2006. doi:10.4135/9780857020116.n192

45. St. George SM, Harkness AR, Rodriguez-Diaz CE, Weinstein ER, Pavia V, Hamilton AB. Applying Rapid Qualitative Analysis for Health Equity: Lessons Learned Using “EARS” With Latino Communities. International Journal of Qualitative Methods. 2023;22:16094069231164938. doi:10.1177/16094069231164938

46. ACHA. Adult Congenital Heart Association. Accessed March 19, 2025. https://www.achaheart.org/

47. Conquering CHD. Conquering Congenital Heart Disease. Accessed March 19, 2025. https://www.conqueringchd.org/

48. The Mended Hearts. The Mended Hearts, Inc. March 19, 2025. https://mendedhearts.org/

49. Team Uncle Joe. Team Uncle Joe: CHD Patient Advocates - Navigating CHD. March 19, 2025. https://teamunclejoe.org/

50. Beatty AL, Peyser ND, Butcher XE, et al. The COVID-19 Citizen Science Study: Protocol for a Longitudinal Digital Health Cohort Study. JMIR research protocols. 2021;10(8):e28169.

51. Marelli A, Rozenblum R, Bolster-Foucault C, et al. Development of MyREADY Transition BBD Mobile App, a Health Intervention Technology Platform, to Improve Care Transition for Youth With Brain-Based Disabilities: User-Centered Design Approach. JMIR Pediatr Parent. 2024;7:e51606. doi:10.2196/51606

52. Nashat H, Habibi H, Heng EL, et al. Patient monitoring and education over a tailored digital application platform for congenital heart disease: A feasibility pilot study. International Journal of Cardiology. 2022;362:68–73. doi:10.1016/j.ijcard.2022.05.002

53. Pew Research Center. Mobile Fact Sheet. Washington. Pew Internet and Technology. November 13, 2024. https://www.pewresearch.org/internet/fact-sheet/mobile/

54. Lopez MH, Gonzales-Barrera A., Patten E M. Closing the digital divide: Latinos and technology adoption. Washington: Pew Internet and American Life Project. 2013;2020(April, 22). http://assets.pewresearch.org/wp-content/uploads/sites/7/2013/03/Latinos_Social_Media_and_Mobile_Tech_03-2013_final.pdf

55. Mora MA, Saarijärvi M, Sparud-Lundin C, Moons P, Bratt EL. Empowering young persons with congenital heart disease: Using intervention mapping to develop a transition program-the STEPSTONES project. Journal of pediatric nursing. 2020;50:e8–e17.

56. Damschroder LJ, Reardon CM, Widerquist MAO, Lowery J. Conceptualizing outcomes for use with the Consolidated Framework for Implementation Research (CFIR): the CFIR Outcomes Addendum. Implementation science. 2022;17(1):1–10.

57. Kuhl EA, Sears SF, Conti JB. Internet-based behavioral change and psychosocial care for patients with cardiovascular disease: a review of cardiac disease-specific applications. Heart & Lung. 2006;35(6):374–382.

58. Klonoff DC. Behavioral theory: the missing ingredient for digital health tools to change behavior and increase adherence. Journal of diabetes science and technology. 2019;13(2):276–281.

