## Supplemental File for "Digital Health Intervention to Promote Lifelong Specialized Care in Adults with Congenital Heart Disease: Study Design"

**Supplementary Online Content**

1. **Semi-structured Interview Guide- Patients**
2. **Semi-structured Interview Guide- Clinicians**
3. **eTable 1: COM-B Targets to CHD Patient Activation and Engagement**
4. **eTable 2: Emerging Hopes for the Digital Tool Prototype**
5. **eTable 3: EmpowerMyCH Surveys**
6. **eTable 4: Exhibit Content Areas and Objectives**
7. **eTable 5: EmpowerMyCH Design Guide**
1. **Semi-structured Interview Guide- Patients**

| **Questions** | **Prompts** |
| --- | --- |
| *Childhood Memories*  Growing up, what was it like to have a heart condition? | - What did you know or understand about your heart condition? How did you gain that knowledge? - What was your understanding about the need for follow up for your heart condition? - What was your understanding about any complications or problems related to your heart in the long-term? |
| *Adult Transition*  Now that you are an adult, what is it to like to have a heart condition? | - How did your knowledge of your heart condition change from when you were a child to when you were an adult? Why do you think this change occurred? - What did you understand or know about how your heart condition will be managed after adulthood? - Looking back at your own care – what would you have liked to know? What would have helped you? What would you have wanted? - What was the transition like between your child and adult heart doctor? - What were your thoughts or feelings around moving from a pediatric to adult doctor? - Have you used any resources to aid your transition to adult care? Why/why not? |
| *Navigating Health System*  What was your experience with the healthcare system like once you were an adult? | - Were there any times when it was [easy OR difficult] to make an appointment? What was that like? - Why do you think it was [easy OR challenging]? - What do you wish you knew beforehand that could have helped you? - How was your experience making an appointment with the adult health care system? - How did you decide who was the right heart doctor for you to make an appointment with? |
| *Gaps in Care*  How do you think your follow-up care for your heart disease has been throughout your adulthood? | - Tell me about your experience when you were not following up with any heart doctor regularly. What was it like? - How long was that time period? - Why do you think you experienced these gaps in care with your heart doctor? - What could have helped you stay in care? - [For no gaps in care] Why do think your follow-up care was consistent? |
| *Benefits/Consequences/Motivations of Being “In Care”*  Seeing a specialist regularly can be challenging on our schedules—what are your thoughts on the need for ongoing care? How do you manage it with your schedule? | - How often do you think someone with your condition should see their heart doctor? Who should be involved in that decision? - How do you manage visiting an ACHD specialist with your other commitments? - What do you think are the biggest challenges? How do you overcome them? - What helped or could help you feel more confident in your abilities to find specialists and schedule meetings with them? - How do you remember to schedule and make an appointment? How do you feel this system is working for you? - What could an ACHD specialist, or another member of your care team, do to make it easier to keep track of appointments? |
| *Intervention Components*  We are going to change topics slightly to talk about potential goals for your healthcare. What are your thoughts about setting some personal goals regarding your health? | - How often do you set goals in your life? What kind of goals do you set? And how do you set them? - How do you work toward achieving those goals? - How do you stay on track to meet your goals? What kinds of things help you measure your progress? - How often do you involve other people in creating or maintaining your goals? Why do you think that is? What [has OR has not] been helpful about having another person involved? What kind of support would you want from another person? - When you think of times where you didn’t reach a goal you wanted, what got in your way? - When have you felt especially motivated to achieve a goal? Where did the motivation come from and what helped maintain it? - [If participant doesn’t set goals] What do you do to achieve the outcomes you want? How has this process worked for you in the past? What do you think could make your process even more useful? |
| *App-based Solutions*  Now, I’d like to focus on some proposed technology-based solutions to possible problems patients face when navigating the healthcare system as an adult.  What are your thoughts about receiving messages through an app on a phone that can support the process of establishing care with your heart doctor? | - What are the kinds of health-related things for which you use an app on the phone? (e.g. exercise tracking, heart rate, etc.) - Can you list or show some of the common apps you use and what you use them for? - How do you see a mobile Health app being able to help you with maintaining life-long care for your heart? - What challenges could you anticipate in having an app like this? What suggestions do you have in order to make this tool easier for you? |

We’re at the end of the interview, but before we conclude I would like to hear about anything you feel we missed.

- Is there anything you’d like to tell me, that maybe I didn't ask you about, but you think it is important for me to know?
- Do you have anything that you would like to ask me/us?

Thank you for your candid responses to our questions.

2. **Semi-structured Interview Guide- Clinicians**

| **Questions** | **Prompts** |
| --- | --- |
| *Introduction to Role/Experiences*  I would like to hear about your experiences caring for patients who have *recently transferred* to adult CHD care.  *are being transferred - *pediatric providers* | - Can you tell me about your current role and the responsibilities that come with it? - How has your experience been providing [adult/pediatric] services? |
| *The CHD Team*  I would like to hear about your experiences with other clinicians in managing CHD patient care. | - Who are the people you consider to be on your team? Are they adult or pediatric based providers? - How is the communication between the different services? - What similarities or differences do you notice between the pediatric and adult services?   - How do these [similarities/differences] influence 1) how you do your job 2) how you view a patient's transition of care? |
| *Independent Decision Making*  What do you think prepares a patient to make independent decisions around their health care? | - How do you assess whether a patient has those things or that knowledge? - What interaction, if any, do you see between your role and preparing a patient to make independent decisions? - What aspects of your role may make it hard to help build this independence? |
| *Transition Facilitators*  What are services or things in the system that  help youth transition from the peds to adult  service? | - What would you consider essential, that if it were removed it would make transitioning or transferring impossible? - What, if added, would ease transitioning or transferring appropriately? |
| *Transition Barriers*  What services or policies are needed to improve the health care transition process? | - What consumes your time? - What affects families? - What are major barriers to transitioning and transfer? |
| *Points for Intervention*  What do you think could be improved with the transfer process from pediatric to adult  focused care or in the process of making patients more independent in making their health-related decisions? | - Are there issues to be solved with the subspecialty services? - Are there issues to be resolved with the outpatient primary care services? - Are there issues to be resolved with the outpatient subspecialty services? - How about community-based services, or other ancillary services? (PT/OT, etc.) |
| *Intervention Components*  What can help patients feel more engaged with their healthcare? | - Are there any tools or resources that could be helpful? - What and when would be the most appropriate way to deliver those resources? - How are your thoughts about patients using a mobile app? - What features of the app would be most useful? - What resources are available and can be used to help patients via app? |

| **eTable 1: Emerging Hopes for the Digital Tool Prototype** | |
| --- | --- |
| **Themes** | **Supporting Narratives** |
| Easy access to credible resources | “I think any kind of connection or links that actually get you to advocacy, or insurance companies, or softwares. Or at least kind of give information on how to navigate those things maybe in a step-by-step way with resources for that.” – Patient, Woman in their 30’s  “just being instructed on how to search for the right healthcare, that went a long way. Versus having to search it up on Google.” - Patient, Man in their 20’s  “I’ve got questions, millions of questions. And it doesn’t seem like there’s anyone that can answer them. It doesn’t feel like there’s any... At least, no one’s told me any resources that I can reach out to” - Patient, Woman in their 30’s  “it’s hard for me to understand: am I doing well or not doing well? Right? I have my annual visits with my cardiologist. Outside of that, I really don’t have a lot of other insights.” – Patient, Man in their 50’s |
| Uplifting of patient voices | “community building within the congenital heart disease community, like being able to talk to other patients. I can help younger patients who maybe are in college or went through what I had to go through in college, help them find care or also look for older patients and just find mentors.” - Patient, Woman in their 20’s  “having a community board or something, where it's like, "I was diagnosed with ASD at this point, and my favorite resource is" – that's where people, I think, connect. Everything is about human connection. “ - Patient, Woman in their 30’s  “And I think that’s what we’ve seen in our advocacy work is I think when you combine patients and doctors, you get the full experience….So, I think if your app. offered that, that would be intriguing of, “Oh, wow. Okay. I’m getting it from every point of view.” - Patient, Woman in their 20’s |
| Customization to patient needs | “I think an app would be very helpful in sharing awareness [of]... the range of CHD symptoms because we group CHD a lot, as we should, but then there’s also that CHD that exists on a spectrum.” - Patient, Man in their 20’s  “I would love to have a drawing of my condition. So, that way, if I went to another doctor, I’m like, “Here. This is what my heart looks like.” It would be good to have like a full list of here’s all your conditions. Here’s all the meds you’re on. So, it would be almost like your medical health record, essentially, like right there in your phone would be really helpful to have. But I think something specifically for your heart condition would be really, really nice.” - Patient, Woman in their 30’s  “I think what's gonna make things more successful is very individualized and localized types of tools.” - Clinician |
| Centering positivity and joy | “It’s colorful and it’s positive.” - Patient, Woman in their 50’s  “I'm envisioning something that's bright and friendly” - Patient, Woman in their 20’s  “maybe having something dedicated to CHD [meet ups]. You know, ‘anybody want to grab a drink or a coffee or whatever and talk?’... like if you sign up for the app, you can see other CHD patients around you, and maybe meet up,” - Patient, Woman in their 30’s |

**eFigure 1: Digital Tool Branding Prototypes**

**Four logo and name combinations developed through iterative community advisory board discussions. Option B (EMPOWER my CONGENITAL HEART) with logo #2 was selected through CAB voting process for its empowering tone and upward arrow symbolizing patient activation.**

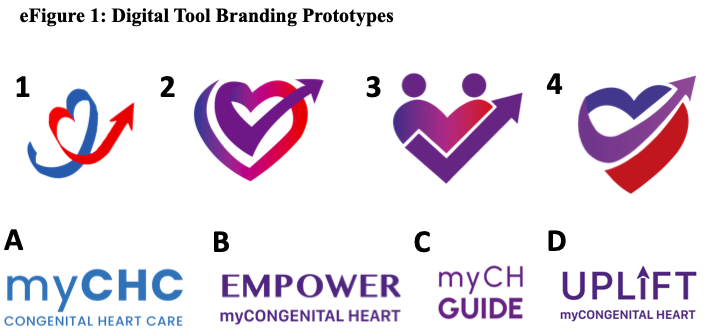

| **eTable 2: EmpowerMyCH Surveys** | |
| --- | --- |
| **Domain** | **Individual Variables** |
| Participants’ Data | |
| Demographics | Age, sex, race/ethnicity, educational status, household income (as a measure of SES), occupation, marital and parental status |
| Diagnosis | Self-reported CHD condition and surgeries, comorbidities |
| Service Use | Outpatient primary care and cardiologist visits, emergency care use or hospitalizations |
| Patient-reported outcomes (PROs) | Euro QoL, Visual Analogue Scale, Health behaviors survey, Illness Identity Survey, self-reported NYHA, Generalized anxiety disorder, PHQ-8, Social history |
| COM-B Patient engagement domains | Patient Activation Measure (13-item, score 0-100; Cronbach’s alpha 0.91)  Gothenburg Empowerment Scale (15-item, score 15-75) |
| Participants’ Feedback | |
| Intervention characteristics | Client satisfaction survey-8 (8 to 32, ~5 minutes, > 90% usual completion rate)  Feedback on each peer and expert advice  Regular app data download to assess use and engagement with various intervention components |
| Participants’ Personal Stories or questions | |
| Patient engagement survey | Opportunity for participants to ask questions or share their insights about managing CHD or connecting with peers. |

| **eTable 3: Exhibit Content Areas and Objectives** | |
| --- | --- |
| **Content Objective** | **Content Area** |
| I know why I should consistently see a provider for my care. | **Getting Started (renamed later as “Finding My ACHD Team”)** |
| I can distinguish between a cardiologist and an ACHD specialist. |  |
| I can find an ACHD provider. |  |
| I know how often I should be seeing an ACHD provider. |  |
| I know how to make an appointment with an ACHD provider. |  |
| I can identify a case manager, or other clinic staff, for questions or concerns related to my upcoming visit. |  |
| I have a system to remind myself when my appointments are. | **Preparing for My Visit** |
| I know what to expect out of my visit. |  |
| I have a system to remember what I want to tell my doctor and what I want to ask my doctor. |  |
| I know to have a paper or electronic file for my medical information. |  |
| I know to ask my doctor or nurse for any recommendations |  |
| I can name and/or describe my heart condition. | **Knowledge about My Heart Condition** |
| I can name and/or describe the cardiac surgeries or procedures I have had. |  |
| I know the names and doses of my medications and when to take them. |  |
| I know what each of my prescribed medications do. |  |
| I can tell whether I need to go to the doctor or whether I can take care of a health problem myself. |  |
| I understand the long-term potential issues associated with my heart condition. |  |
| I know what my typical vital signs are and have a copy of my ECG. |  |
| I understand the possible side effects associated with my medications. |  |
| I know if and when I should take antibiotics prior to dental procedures. |  |
| I know about medications or supplements that could interfere with my heart medications. |  |
| I know what types of activities or exercises are safe and healthy for me to do. |  |
| I understand the impact of tobacco, alcohol, recreational drug use, and unprotected sex on my heart and health overall. |  |
| I understand the risk of passing on my heart condition to future children. |  |
| I understand the risk of pregnancy and the need for pre-pregnancy counseling. |  |
| I understand what types of contraception are safe for me based on my heart condition. |  |
| I know why I need health insurance. | **Insurance** |
| I understand how health insurance works. |  |
| I know basic health insurance terms. |  |
| I can differentiate between the different kinds of health insurance plans. |  |
| I know where to find health insurance. |  |
| I know what type of insurance policy to pick. |  |
| I know how to contact my health insurance with questions or concerns. |  |
| I know what health information to carry with me daily. |  |
| I can explore the essential benefits of health insurance policies. |  |
| I have a plan for how I will get to my appointments. | **Transportation** |
| I understand non-emergency medical transportation options available to me. |  |
| I know whether or not my insurance supports medication delivery from my pharmacy to my home. |  |
| I know how to get to my doctor's office. |  |
| I understand the work benefits related to my heart condition (sick leave, FMLA, short term disability). | **Legal Considerations** |
| I understand the education benefits related to my heart condition (504, IEPs) |  |
| I know what an advance directive is and how to set one up. |  |
| I understand what I need/don't need to tell my employer in relation to my heart condition. |  |
| I know how to get connected with others who have a heart condition like mine. | **Community & Belonging** |
| I am aware of organizations for those who have a heart condition like mine. |  |
| I have someone to motivate me toward my goals. |  |

| **eTable 4: EMCH Design Guide** | |
| --- | --- |
| **Section Statements** | **Supporting Narratives** |
| Messaging statement | To empower CHD patients to proactively participate in their health journey and thrive: Every hero embarks on a journey marked by challenges and uncertainties. For patients with congenital heart disease the journey through the healthcare system can often feel daunting and overwhelming. Just as every hero experiences moments of confusion and struggle, CHD patients often find themselves navigating complex medical landscapes and decisions.  By framing the patient's experience within this heroic narrative, we ensure that our guidance is both empowering and supportive. |
| Design statement | We value **readability** above all else. These design guidelines serve as a foundation for consistency and structure; if a rule compromises the clarity of the content, make exceptions to improve the design. If an element doesn't look correct or detracts from the user experience, adjust the design rather than adhere strictly to the rules. |
| Written language statement | We build our tools around the written word; we have an incredible opportunity to convey our message with precision and impact. Clarity, simplicity, and brevity are essential to ensuring that our message resonates with and reaches the appropriate audience. Every word choice is critical—each must be deliberate, contributing to a message that is both powerful and **accessible**. We prioritize **readability** in both our language and design—no amount of design can compensate for poorly crafted language. To support this, we can use tools like Simple Measure of Gobbledygook ([SMOG](https://www.ncbi.nlm.nih.gov/pmc/articles/PMC5764592/)), which estimates the readability of text by determining the grade level needed to understand it based on the frequency of polysyllabic words. By focusing on the craft of language, we can balance style and substance, leading to consistently clear and effective communication that can meet these readability standards. |
